# Evaluation of Covid-19 Ag-RDTs self-testing in Lesotho and Zambia

**DOI:** 10.1101/2022.12.21.22283827

**Authors:** M. Bresser, R.M. Erhardt, K. Shanaube, M. Simwinga, P.A. Mahlatsi, J. Belus, A. Schaap, A. Amstutz, T. Gachie, T.R. Glass, B. Kangolo, M.J. ‘Mota, S. Floyd, B. Katende, E. Klinkenberg, H. Ayles, K. Reither, M. Ruperez

**Author notes:** These two authors contributed equally to this work (M. Bresser, R.M. Erhardt). Klinkenberg, E. conducted this work as consultant for LSHTM.

## Abstract

**Background:** The use of antigen rapid tests (Ag-RDTs) for self-testing is an important element of the COVID-19 control strategy and has been widely supported. However, scale-up of self-testing for COVID-19 in sub-Saharan Africa is still insufficient and there is limited evidence on the acceptability of self-testing and agreement between Ag-RDT self-testing and Ag-RDT testing by professional users. A joint collaboration (BRCCH-EDCTP COVID-19 Initiative) was established between Lesotho and Zambia to address these gaps in relation to Ag-RDT self-testing and contribute to increasing its use in the region.

**Methods:** A cross-sectional study was conducted with qualitative and quantitative data analysis. Firstly, 11 in-depth cognitive interviews (5 in Zambia and 9 in Lesotho) were performed to assess the participants’ understanding of the instructions for use (IFU) for self-testing. In a second step, evaluation of test agreement between Ag-RDT self-testing and Ag-RDT testing by professional user using SD Biosensor STANDARD Q COVID-19 Ag-RDT was performed. In Zambia, usability and acceptability of self-testing were also assessed.

**Results:** Cognitive interviews in Lesotho and Zambia showed overall good understanding of IFU. In Zambia, acceptability of self-testing was high, though some participants had difficulties in conducting certain steps in the IFU correctly. Agreement between Ag-RDT self-test and Ag-RDT by professional users in Lesotho (428 participants) and Zambia (1136 participants) was high, 97.6% (404/414, 95% CI: 95.6-99.8) and 99.8% (1116/1118, 95% CI: 99.4-100) respectively.

**Conclusion:** Findings from this study support the use of Ag-RDT self-testing within COVID-19 control strategies in sub-Saharan Africa, contributing to increase the testing capacity and access in hard-to reach settings.

## Introduction

Since the emergence of SARS-CoV-2 in December 2019 and the declaration of the pandemic by the World Health Organization in March 2020, 640 million Covid cases and more than 6.6 million deaths worldwide have been reported (1). In sub-Saharan Africa (SSA), this accounted for about 7 million reported Covid-19 cases and 175,000 deaths (1). Many infections have gone undetected; SSA has one of the highest estimates of excess mortality versus reported mortality ratios (2). To scale up detection of COVID-19 infections, recommended control strategies from national and international bodies such as the WHO and the African Centres for Disease Control and Prevention (Africa CDC) have shifted from nucleic acid amplification tests to antigen rapid diagnostic tests (Ag-RDT) and from testing by health care workers to self-testing (3, 4). However, the scale-up of Ag-RDT COVID-19 self-testing in the African continent is still insufficient.

Ag-RDT self-testing as a potential testing strategy appears both feasible (5-8) and acceptable (8-14). However, most of the evidence comes from high- and middle-income settings and there is very little evidence from SSA or other low and middle income areas(3, 4). Results from other disease areas however, such as Human Immunodeficiency Virus (HIV), showed implementation of self-tests in low- and middle-income countries (LMICs) is both feasible and acceptable (7), pointing to the potential of Ag-RDT self-testing. The question of whether Ag-RDT testing can be shifted from health care workers to self-testing by the general population is particularly important in less urban areas, where educational level, health literacy and literacy rates tend to be lower (15-17). This could potentially limit understanding of self-test instructions and hence limit ability to perform self-testing accurately (18). If self-testing in SSA can produce similar results as with testing by health care workers, this could alleviate the burden on health systems and interference with other diseases service provision during future SARS-CoV-2 waves through cost reductions on staff for testing or by reassignment of staff to other pandemic control tasks. Moreover, appropriate self-test distribution programs could drastically increase access to testing and reduce the response time to implement control measures limiting transmission into the community.

The current interim guidances on COVID-19 antigen self-testing released by the WHO and Africa CDC that recommend incorporating self-testing into national testing strategies, are based mainly on studies from high income countries and urban settings (3, 4). These studies furthermore vary considerably in study design (e.g. sample type, reference standard, population), making it difficult to test for operator effects; if the ability to perform the self-test depends on user characteristics such as literacy, health literacy and previous testing experience. There is evidence from studies in high income settings assessing performance of self-sampling tested against Polymerase Chain Reaction (PCR), with reported yields close to or over the WHO sensitivity and specificity thresholds of 80% and 97% respectively (19-24). It is necessary to expand on these findings and assess whether both self-collection of samples and self-testing can be accurately performed by the general population in LMICs. This includes all steps related to testing; from swab collection to processing and accurately interpreting the result.

A joint COVID-19 BRCCH-EDCTP Collaboration Initiative aimed to address the research gaps on Ag-RDT self-testing in hard-to-reach areas and vulnerable populations and to build up existing knowledge surrounding self-testing by assessing the usability and acceptability of Ag-RDT self-testing and agreement with testing by health care workers in Lesotho and Zambia.

## Methods

### Study design

This is a cross-sectional study with analysis of both qualitative and quantitative data, evaluating i). participant understanding of Instruction for Use (IFU) for COVID-19 Ag-RDT self-testing using cognitive interviews in Lesotho and Zambia, ii). ease-of-use of Ag-RDTs for self-testing in Zambia using a standardized questionnaire, iii) usability of Ag-RDT self-testing through observation and assessment of participants conducting all the steps in the IFU in Zambia and iv). level of agreement between self-testing and testing by a professional user performing/conducting Ag-RDTs for detecting COVID-19 infection in Lesotho and Zambia.

In both countries, this study was nested within larger community-led projects. In Zambia, this study was conducted as part of the TREATS-COVID study that aimed at measuring the prevalence and spread of SARS-CoV-2 in the community, whereas in Lesotho it was nested within the MistraL project, which assessed the effects of different interventions on mitigating COVID-19 pandemic in the community.

### Study population

In Zambia, this study was conducted in a middle- to high-density peri-urban area location about 15 km north-west of Kabwe town center in Central Province, Zambia. HIV prevalence in this community is approximately 15% and tuberculosis (TB) prevalence is estimated to be in the range of 0.5%-1%. The total population is around 28,000 individuals, living in approximately 5,300 households (average household size around 5.3), of whom around 17,000 (60%) are aged ≥15 years.

In Lesotho, this study was conducted in 23 villages in the municipality of Butha-Buthe, located in both semi-urban and rural areas (9 peri-urban, 14 rural). HIV prevalence rate in this community is approximately 17.8% as of 2017 (25) and TB prevalence rate in Lesotho is estimated to be 531/100,000 and 670/100,000 for peri-urban and rural areas, respectively. Village size of people aged ≥18 years ranges from 40 to 303, adding up to a total population of ca. 40,000.

### Study procedures

#### Evaluation of the understanding of manufacturer’s IFU for self-testing by participants in Lesotho and Zambia using cognitive interviews

In Zambia as well as Lesotho, cognitive interviews were conducted with potential participants to explore their understanding of the manufacturers’ IFU. A convenience sampling approach was used at both sites. In Zambia, participants (three males and two females) were selected from among those who visited community centers for testing to represent intended users of the COVID-19 Ag-RDT self-test kits. The selected participants were also required to complete a simple functional test that required them to read a few lines of the consent form in English. In Lesotho, community health workers (CHWs) contacted nine participants (five males and four females) who they knew were symptomatic and represented potential users of the COVID-19 Ag-RDT self-test kits.

In both countries, cognitive interviews were conducted using an in-depth interview (IDI) guide structured according to the steps in the manufacturer’s IFU of the SD Biosensor STANDARD Q COVID-19 (SD Biosensor; Republic of Korea) Ag-RDT kit.

Participants were given an Ag-RDT self-test kit, which contained the manufacturer’s IFU. In Zambia, a trained social scientist probed participants’ cognition of the different steps in the IFU. For every step on the IFU, participants were asked to view the pictorial instructions (pictures), read the written instructions, reflect on what they were instructing them to do and say this loudly. Participants were subsequently asked to perform all the actions without assistance. Differently to Zambia, in Lesotho, in-depth interviews were conducted by trained research assistants after participants had self-tested following the manufacturer’s IFU. In both countries, participants were asked to reflect on whether other members of the community would understand the instructions in the IFU and correctly perform the actions. All interviews were audio recorded.

In Lesotho, the findings from cognitive interviews were used to make adaptations to the IFU that was later used in the evaluation of test agreement between self-testing and testing by CHW

#### Evaluation of ease-of use, usability and agreement between self-testing vs testing by CHW

An overview of the study procedures in Lesotho and Zambia is displayed in table 1.

**Table 1.**
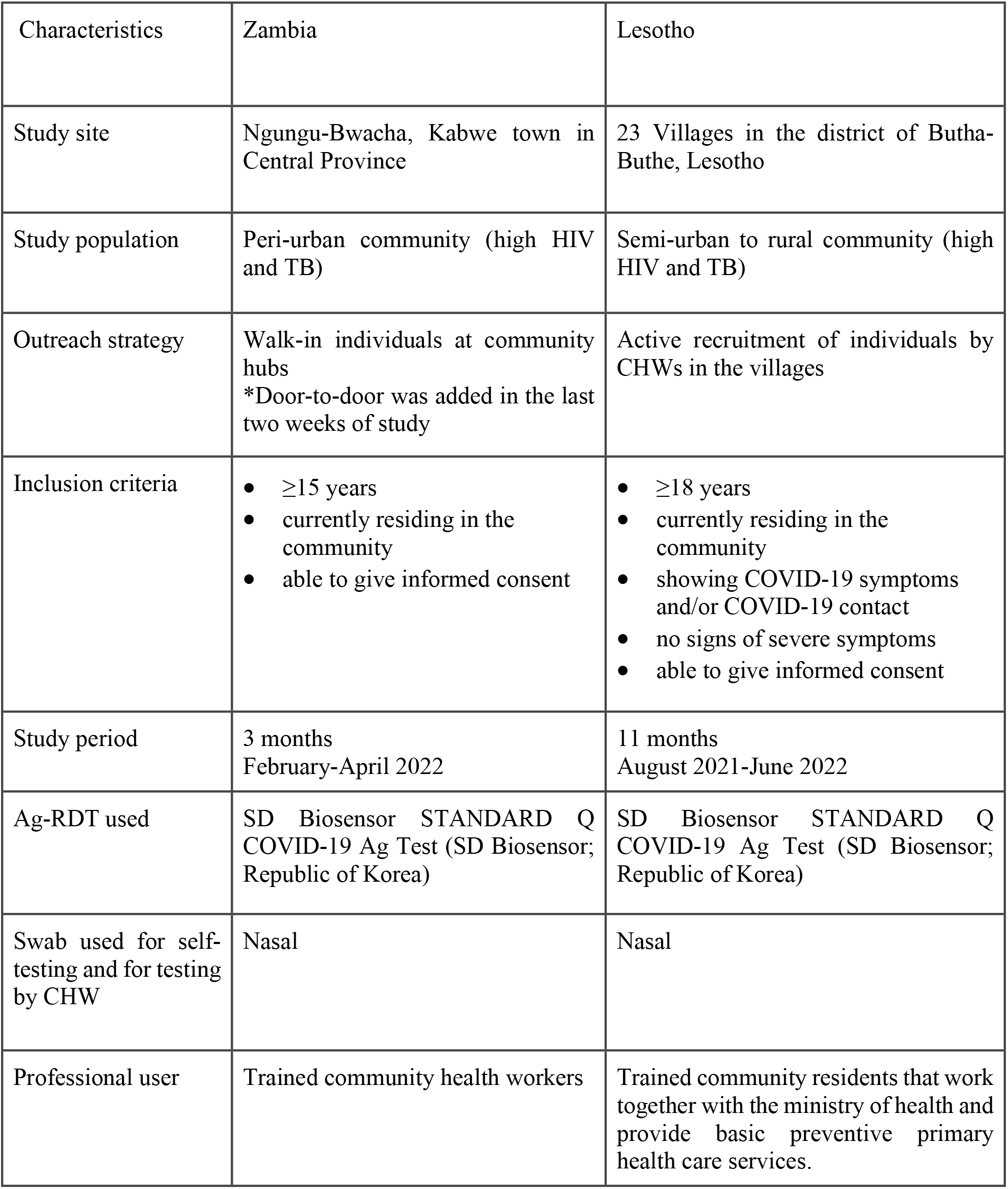
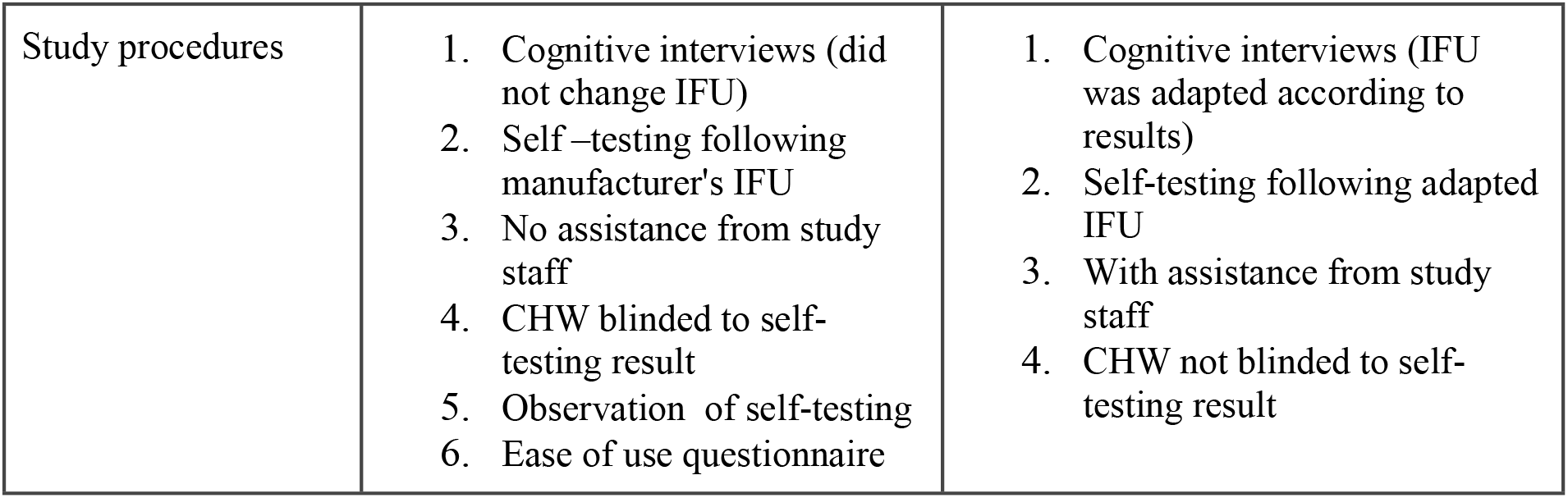
Outline of study characteristics in Zambia and Lesotho.

In Zambia, walk-in testing locations called community hubs were set up at different strategic points in the community. Community hubs were staffed by CHWs and had a flexible set-up, allowing them to be moved around in the community. Regular sensitization and awareness campaigns were conducted in specific high-risk transmission hotspots in the community to encourage the population to attend the hubs.

At the community hubs, walk-in individuals who were eligible to participate in the study (>=15 years, residing in the community and able to give informed consent) were administered a questionnaire on socio-demographic characteristics and COVID-19 suggestive symptoms and were offered to self-test using the SD Biosensor STANDARD Q COVID-19 Ag-rapid-test and to be tested by a CHW using the same test. Half-way through the study, in order to increase the proportion of women enrolling, some CHWs also went door-to-door in the community offering eligible household members to participate and get self-tested and tested by CHWs on-the-spot, at their households.

Individuals were given a self-test kit containing the manufacturer’s instructions for use (IFU) (S1 Appendix), a nasal swab, the testing device and a form to fill in results and ease-of-use questions (S2 Appendix). Participants were asked to read and follow the manufacturer’s IFU, self-collect a nasal sample, perform the test and fill in the form without any interference or assistance by study staff. During the self-testing process, participants were observed by a designated observer who completed an observational form if key steps according to the manufacturer’s IFU were conducted appropriately by the participant. The observer was not allowed to assist or interact with the participant during the process. If the result from the self-test was ‘invalid’, according to the manufacturer’s IFU the self-test had to be repeated. Once the self-testing process was completed, the observer compared the result on the test device with the results recorded by the participant on the form, and documented any inconsistencies between the two.

Directly following the self-testing, the participant was tested by the CHW, who was blinded to the self-testing results. The CHW user collected a nasal sample from the participant, conducted the Ag-RDT and recorded the result in the Electronic Data Capture tool. If the result from the test was ‘invalid’, another test was performed with the remaining sample in the extraction tube. If there was not enough sample or if the result was ‘invalid’ again a new nasal swab was collected from the participant and tested again.

A participant was classified as being COVID-19 positive if any of the two tests, the self-test or the test performed by the CHW was positive, whereas participants were classified as being COVID-19 negative if both tests were negative.

In Lesotho, participants were recruited by CHWs; community residents that work together with the Ministry of Health and provide basic preventive primary health care services, as well as screening for various health problems. Outreach took place within the community, with individuals who were symptomatic or had contact with a COVID-19-positive case being approached by the CHW.

Individuals who were eligible (≥18 years, residing in the community and able to give consent, COVID-19 contact or symptomatic and agreed to participate in the study) were provided with a self-test kit, SD Biosensor STANDARD Q COVID-19 Ag Test, which included the adapted IFU, the swab, and the test device. Participants were guided by the CHW through all steps of the adapted IFU (S3 Appendix). After the self-test procedure was explained by the CHW, participants were asked to self-collect a nasal sample and perform the Ag-RDT themselves. However, in contrast to Zambia, the CHW was present during all steps, was able to assist if any of the steps were unclear and intervened if participants were not following the IFU correctly. The self-test results were read and entered by the CHW into the e-application or paper form. In case of an invalid test result the test was repeated with a new sample, and the final result was recorded. Once the participants had self-tested, they were tested by the CHW with the same Ag-RDT. The same CHW who prior assisted in the self-testing, collected a nasal sample from the participant, conducted the Ag-RDT and recorded results on the e-application or paper form. In case of an invalid test result the test was repeated. Unlike Zambia, the CHWs in Lesotho were not blinded to the result of the self-test. The participant was considered COVID-19 positive if any of the two Ag-RDT tests was positive.

For both countries, national guidelines were followed in case a participant was COVID-19 positive.

### Data management and statistical analysis

For baseline characteristics, descriptive statistics were used. Dichotomous variables were reported as numbers with percentages whilst continuous variables were reported with the median and interquartile range (IQR). Because different procedures were used, the analysis comparing results of self-testing with those of testing by the CHW was conducted separately for Zambia and Lesotho though using the same measures. We calculated the point estimates and 95% confidence intervals (CIs) for the level of agreement. Participants with missing results for either self-testing or testing by CHW were excluded from this analysis.

Definitions for symptomatic, TB and HIV status, usability and acceptability are detailed in S4 Appendix.

In Zambia analyses were performed using STATA 17, in Lesotho STATA 16 (STATA, USA).

### Ethics

In Zambia, this study was approved by the research ethics committees of the London School of Hygiene & Tropical Medicine and the University of Zambia (UNZABREC) and National Health Research Authority. In Lesotho, this study was approved by the National Health Research Ethics Committee Lesotho.

## Results

### Understanding of the manufacturer’s IFU for self-testing using cognitive interviews in Lesotho and Zambia

The manufacturer’s IFU used in both countries asked the self-tester to check the expiry date and the presence of a desiccant in the package as a first step/check before proceeding. This was followed by instructions on washing or sanitizing one’s hands at the start of the procedure and not touching the tip of the swab when opening the swab pouch, tilting the head at an angle and inserting swab into the nostrils up to a correct depth, and rotating the swab for a specific number of times. Specimen processing instructions included how to apply 4 drops of the specimen collected to the specimen well, waiting for and interpreting the results.

Most of the participants in both countries said they understood the pictures and written instructions in the IFU, the written and pictorial instructions were said to be complementary. Most of the participants indicated they relied on the pictures to perform actions in the IFU. Almost all participants correctly interpreted their test results even though some did not understand the meaning of invalid results. There were some steps in the IFU for which most participants indicated they did not understand both the written and pictorial instructions. An example in Zambia for the instruction that required the participant to set the solution tube on the stand hole. Most participants could not find the area to perforate the package to make the stand hole, but all found a way of ensuring that the solution tube stood upright by setting it against the package.

Participants from both countries observed that some key items were not included with the test kits such as hand sanitizers, watches and gloves, and the need for such items was not mentioned in the first steps of the IFU so that participants could prepare and look for them or their alternatives before proceeding.

Some pictures were not clear. Participants in both countries did not recognize/ understand the image of a clock. “*For people to know it is 15 minutes, you need to put a watch with labels or handles and numbers”* (woman, 66 years, Zambia). Participants also found it difficult to understand pictorial instructions where many pictures were clustered together. Use of difficult words and terms such as ‘nozzle cap’, ‘desiccant’, ‘foil pouch’, ‘specimen well’, ‘sterile swab’, ‘nostril’ also affected participants’ comprehension of the written instructions. One essential information, expiry date, was not easy to find by some participants in Zambia, while all participants in Lesotho easily and quickly found it. None of the participants knew the use of the desiccant. One participant in Zambia thought the desiccant was to be poured (later) in the solution/ buffer tube. All the participants correctly tested and interpreted their results. However, most participants anticipated that other community members, particularly those with low literacy, would struggle to understand the instructions with the current limitations, which could lead to performance errors. As a result of this feedback, the team in Lesotho made minor updates to the IFU used in the main study, the major change being the translation of the IFU into the local Sesotho language, with certain words adapted to the local context. Another change was an adaptation of the images showing the test results.

### Ease-of use and usability of self-testing with Ag-RDTs in Zambia

Table 2 depicts the results from the ease-of-use questionnaire in Zambia. 1125 participants filled in the ease-of-use questionnaire, 86% (962/1125) found that instructions for use were ‘very easy to understand’ and, similarly, 92% (1,032/1125) answered that the Ag-RDT was ‘very easy’ to do. Most of the participants relied on the pictures and indicated that pictures were the most helpful piece of the instructions. Overall, 92% (1038/1125) were ‘very satisfied’ with the self-testing process, 90% (1027/1125) trusted the results obtained and almost all (97%, 1095/1125) would be ‘very likely’ to recommend doing a self-test to a friend.

**Table 2.** ‘Ease of use’ assessment self-completed questionnaire on self-testing with in Zambia (N=1125)

| Questions | Responses | n | % |
| --- | --- | --- | --- |
| **How easy was the COVID Ag-RDT self-test to do?** | Very easy | 1032 | 91.7 |
|  | Somewhat easy | 88 | 7.8 |
|  | Somewhat difficult | 4 | 0.4 |
|  | Very difficult | 1 | 0.1 |
| **Which part of the test did you find the most difficult to perform?** | None of the steps were difficult | 44 | 4.3 |
|  | Putting the buffer into the extraction tube | 7 | 1.0 |
|  | Collecting the nasal swab. | 9 | 1.0 |
|  | Swirling the swab in the extraction tube | 8 | 1.0 |
|  | Dispensing 5 drops from the tube into the test device | 10 | 1.1 |
|  | Reading the test. | 3 | 1.4 |
|  | All the steps were easy | 0 | 0.0 |
|  | Other difficulty | 7 | 0.7 |
| **How easy was it to understand the instructions provided?** | Very easy | 962 | 85.5 |
|  | Somewhat easy | 141 | 12.5 |
|  | Somewhat difficult | 16 | 1.4 |
|  | Very difficult | 6 | 0.5 |
| **How much did you rely on the pictures to conduct the test?** | Very much | 951 | 84.5 |
|  | Somewhat | 47 | 4.1 |
|  | A little | 106 | 9.4 |
|  | Not at all | 21 | 1.9 |
| **Which was the most helpful piece of instruction?** | Pictures | 942 | 83.7 |
|  | Written instructions | 183 | 16.3 |
| **Overall, how satisfied were you with the self-testing process?** | Very satisfied | 1038 | 92.3 |
|  | Somewhat satisfied | 66 | 5.9 |
|  | A little satisfied | 16 | 1.4 |
|  | Not satisfied at all | 5 | 0.4 |
| **Do you believe that your test result was correct?** | Definitely correct | 1017 | 90.4 |
|  | Probably correct | 108 | 9.6 |
|  | Not correct | 0 | 0 |
| **Would you recommend this COVID test kit for self-testing to friends and family?** | Yes | 1113 | 98.9 |
|  | No | 12 | 1.1 |
| **How likely would you be to self-test in the future if kits were made available for the general public?** | Very likely | 1095 | 97.3 |
|  | Somewhat likely | 24 | 2.1 |
|  | Not likely at all | 2 | 0.2 |
|  | Never again | 4 | 0.4 |

Table 3 shows the percentage of participants who were ‘observed’ to conduct correctly each of the different steps listed in the manufacturer’s IFU used in Zambia out of the 1119 participants that went through the observation process. Most of the steps were conducted correctly by more than 85% of participants, except placing the tube on the kit box tray holder, swirling the swab in the fluid 5 times while pushing it against the walls of the tube, removing the swab slowly while squeezing the sides of the tube to extract the liquid from the swab, breaking the swab correctly and pressing the nozzle cap tightly onto the tube which were conducted correctly by 72% (805), 71% (799), 56% (625) and 60% (672) of the participants, respectively.

**Table 3.** Observation checklist on steps listed on instructions for use in Zambia (N=1119)

| **Self-testing steps described in the Instructions for Use** | **n** | **%** |
| --- | --- | --- |
| **Did participant place tube on the kit box tray holder correctly?** | 805 | 71.9 |
| **Did participant insert swab into the left nostril to the correct depth?** | 1106 | 98.8 |
| **Did participant rotate the swab 5 times in the left nostril?** | 1018 | 91.0 |
| **Did participant insert the swab into the right nostril to the correct depth?** | 1094 | 97.8 |
| **Did the participant rotate the swab 5 times in the right nostril?** | 1094 | 97.8 |
| **Did the participant insert the swab into the solution tube correctly?** | 1117 | 99.8 |
| **Did the participant swirl in the fluid 5 times while pushing against the wall of the Tube** | 799 | 71.4 |
| **Did the participant remove the swab slowly while squeezing the sides of the tube to extract the liquid from the swab?** | 625 | 55.9 |
| **Did the participant break the swab correctly and press the nozzle cap tightly onto the tube?** | 672 | 60.1 |
| Did the participant squeeze 5 drops of liquid from the Tube into the well on the Test Device? | 986 | 88.1 |
| Did the participant read the test result in 15 minutes? | 1113 | 99.5 |

### Evaluation of the agreement between self-testing and testing by CHW in Lesotho and Zambia

#### Baseline characteristics of participants

Baseline characteristics of participants are displayed in table 4. In Zambia, 1137 participants were enrolled from February to April 2022 at community hubs while in Lesotho 428 participants were enrolled from August 2021 to July 2022 from households. In Zambia 38% (432/1137) were females while in Lesotho this was 64% (275/428). Median age of participants was 27 (IQR: 21-37) and 43 (IQR: 31-59) years in Zambia and Lesotho, respectively. Proportion of people who self-reported to be on antiretroviral therapy was 19.4% (83/428) in Lesotho. In Zambia, 4.4% (50/1137) were PLHIV among whom 41 self-reported to be positive and 9 were newly diagnosed at the study. From the remaining 1087, 28 had an unknown status, 544 self-reported to be negative and were not tested again and 515 were negative when tested within the study. The proportion of participants on TB treatment at the time of the study was low in both countries (0.4%).

**Table 4.**
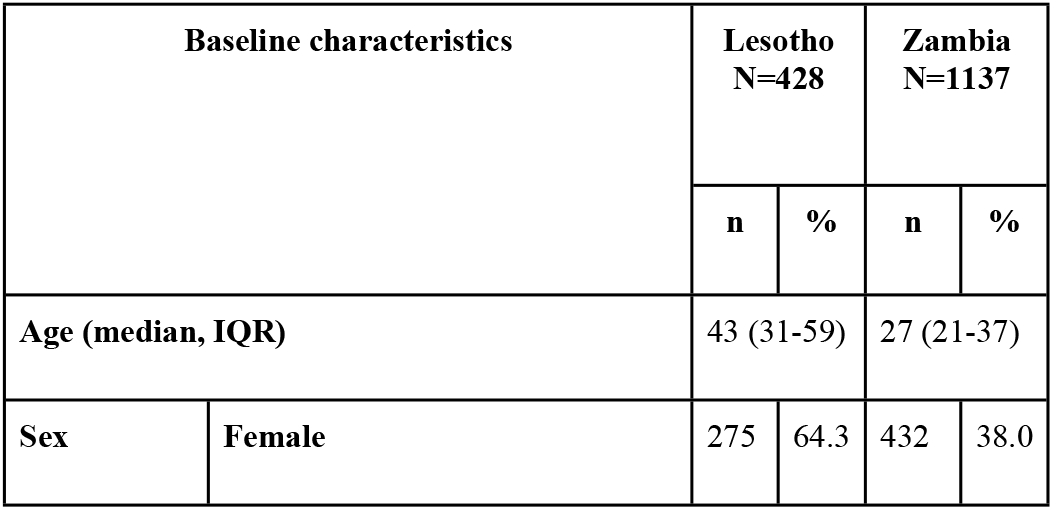
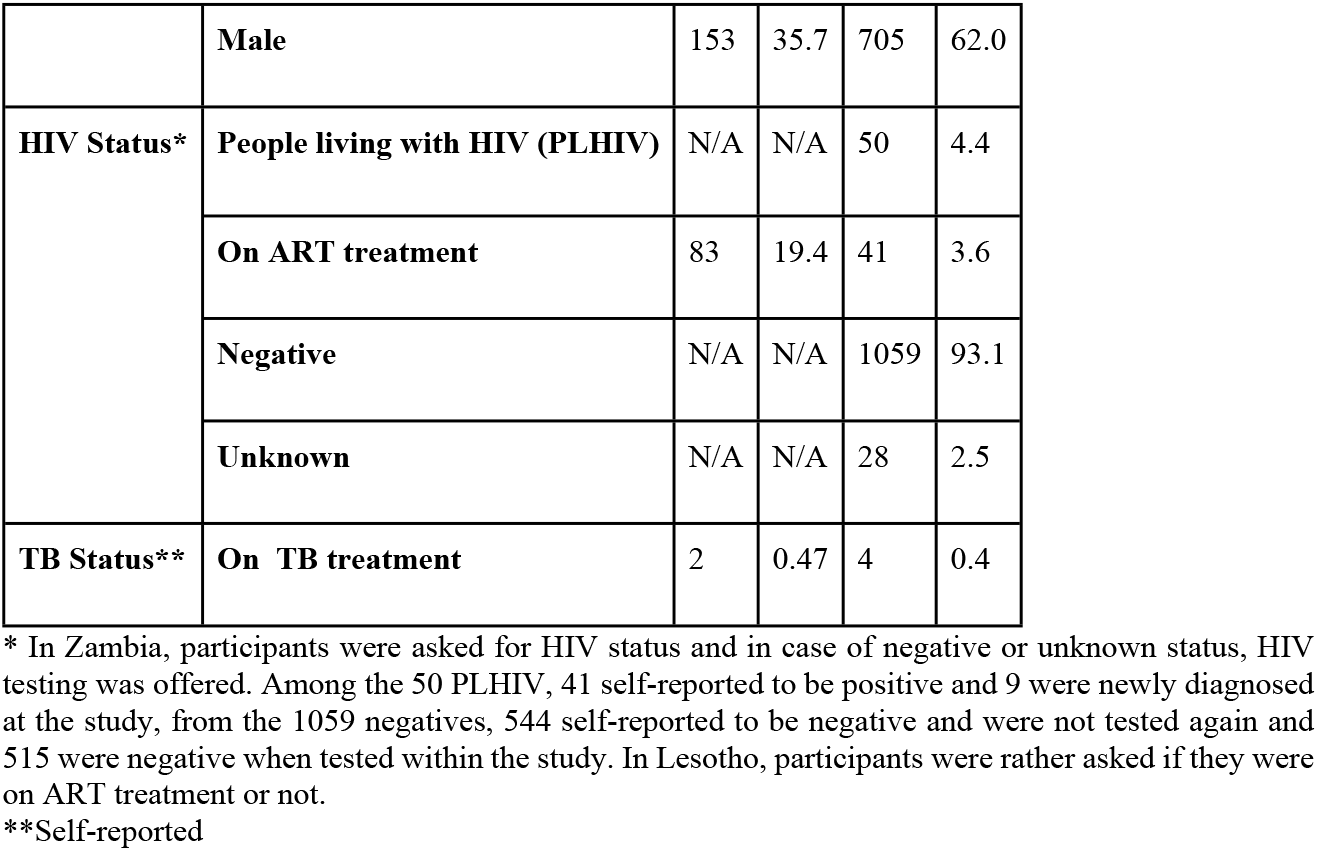
Baseline characteristics of study population in Lesotho and Zambia.

| Baseline characteristics |  | Lesotho<br>N=428 |  | Zambia<br>N=1137 |  |
| --- | --- | --- | --- | --- | --- |
|  |  | n | % | n | % |
| Age (median, IQR) |  | 43 (31-59) |  | 27 (21-37) |  |
| Sex | Female | 275 | 64.3 | 432 | 38.0 |
|  | **Male** | 153 | 35.7 | 705 | 62.0 |
| **HIV Status\*** | **People living with HIV (PLHIV)** | N/A | N/A | 50 | 4.4 |
|  | **On ART treatment** | 83 | 19.4 | 41 | 3.6 |
|  | **Negative** | N/A | N/A | 1059 | 93.1 |
|  | **Unknown** | N/A | N/A | 28 | 2.5 |
| **TB Status\*\*** | **On TB treatment** | 2 | 0.47 | 4 | 0.4 |

\* In Zambia, participants were asked for HIV status and in case of negative or unknown status, HIV testing was offered. Among the 50 PLHIV, 41 self-reported to be positive and 9 were newly diagnosed at the study, from the 1059 negatives, 544 self-reported to be negative and were not tested again and 515 were negative when tested within the study. In Lesotho, participants were rather asked if they were on ART treatment or not.
\*\*Self-reported

#### Symptoms and duration of symptoms in study participants

Table 5 depicts symptoms and symptom duration as reported by participants. Among participants in Zambia, 15% (199/1136) were symptomatic whereas in Lesotho this proportion was much higher, 62% (262/426). The most common symptom reported in both countries among participants with symptoms was cough (74%, 147/199 in Zambia, 70%, 183/262 in Lesotho) followed by chest pain in Zambia (25%, 49/199) and fever in Lesotho (14%, 36/262). In both countries most of the participants had been symptomatic for 7 days or less (94% in Lesotho and 92% in Zambia) and more than half for 3 days or less (64% in Lesotho and 55% in Zambia).

**Table 5.**
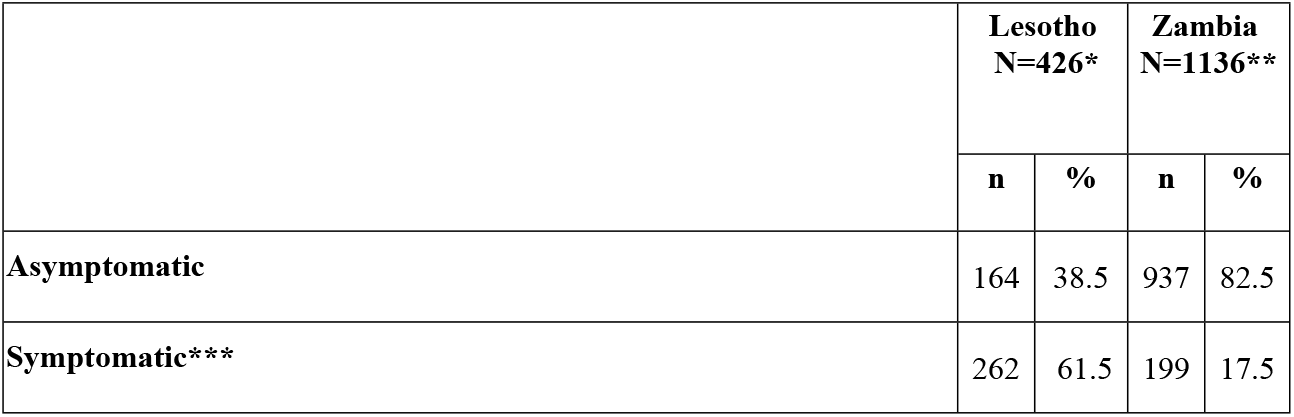
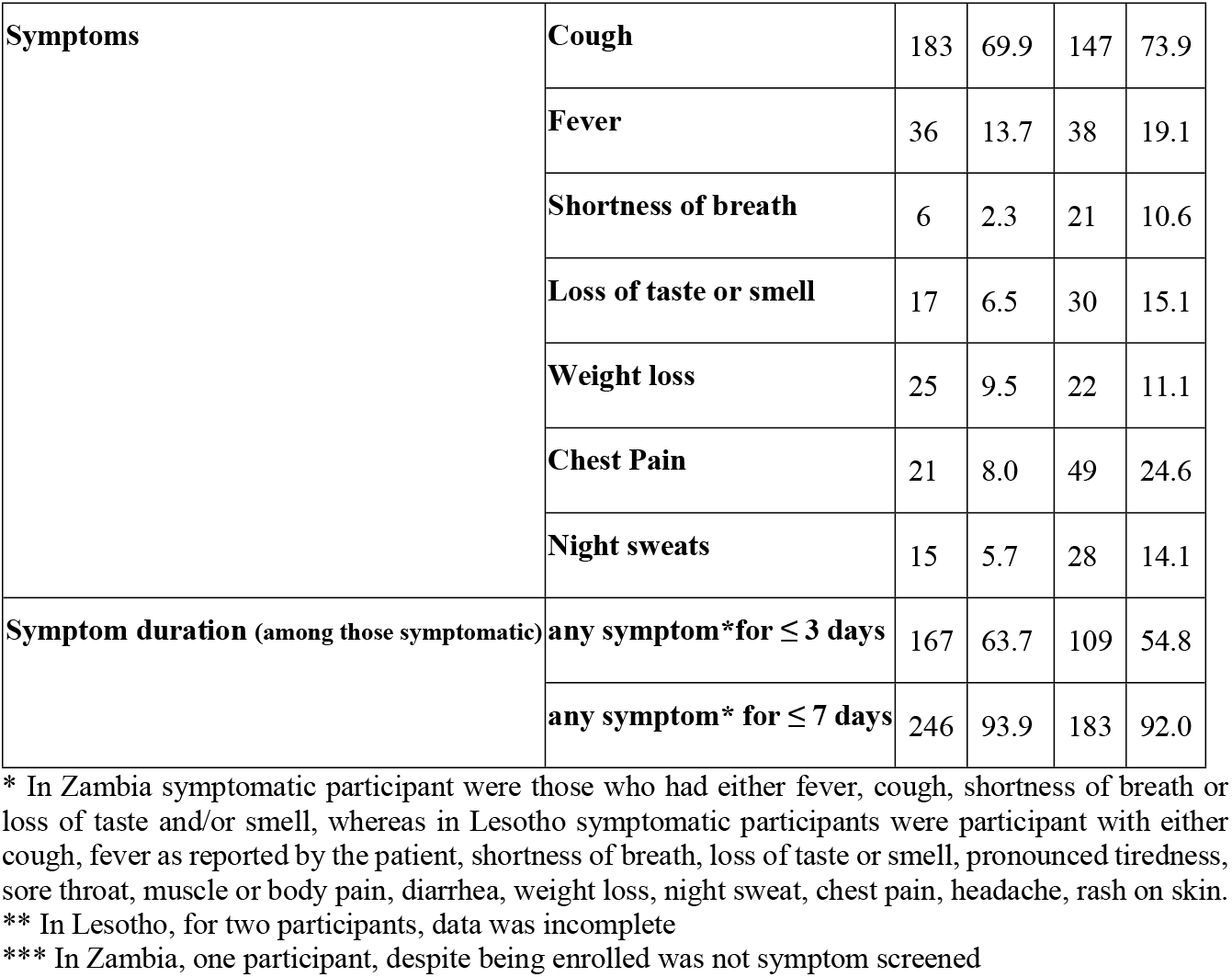
COVID-19 related symptoms and duration in study participants in Lesotho and Zambia.

|  | **Lesotho**<br>**N=426\*** |  | **Zambia**<br>**N=1136\*\*** |  |
| --- | --- | --- | --- | --- |
|  | **n** | **%** | **n** | **%** |
| **Asymptomatic** | 164 | 38.5 | 937 | 82.5 |
| **Symptomatic\*\*\*** | 262 | 61.5 | 199 | 17.5 |

|  |  |  |  |  |  |
| --- | --- | --- | --- | --- | --- |
| **Symptoms** | **Cough** | 183 | 69.9 | 147 | 73.9 |
|  | **Fever** | 36 | 13.7 | 38 | 19.1 |
|  | **Shortness of breath** | 6 | 2.3 | 21 | 10.6 |
|  | **Loss of taste or smell** | 17 | 6.5 | 30 | 15.1 |
|  | **Weight loss** | 25 | 9.5 | 22 | 11.1 |
|  | **Chest Pain** | 21 | 8.0 | 49 | 24.6 |
|  | **Night sweats** | 15 | 5.7 | 28 | 14.1 |
| **Symptom duration (among those symptomatic)** | **any symptom\*for $\leq 3$ days** | 167 | 63.7 | 109 | 54.8 |
|  | **any symptom\* for $\leq 7$ days** | 246 | 93.9 | 183 | 92.0 |

\* In Zambia symptomatic participant were those who had either fever, cough, shortness of breath or loss of taste and/or smell, whereas in Lesotho symptomatic participants were participant with either cough, fever as reported by the patient, shortness of breath, loss of taste or smell, pronounced tiredness, sore throat, muscle or body pain, diarrhea, weight loss, night sweat, chest pain, headache, rash on skin.
\*\* In Lesotho, for two participants, data was incomplete
\*\*\* In Zambia, one participant, despite being enrolled was not symptom screened

#### Test agreement between Ag-RDT self-testing and Ag-RDT testing by CHW

In Zambia, from the 1136 participants that were screened for symptoms, 14 (1%) refused to either self-test and/or be tested by CHW. Of the 1123 participants who had both tests done, 9 participants had a positive result and 1107 a negative one in the self-testing and when tested by the CHW. Five participants had an invalid result when they self-tested but when they were tested by professional user, 4 participants had a negative result and 1 a positive one.

In Lesotho among the 428 participants who were eligible, only 2 refused the self-test and/or the CHW-taken test, the remaining 15 were not tested due to other reasons. From the remaining 413, 38 had a positive result on the self-test, out of whom 37 also tested positive by CHW. From the 375 participants with negative results on the self-test, 9 tested positive by the CHW.

In table 6, results are presented for percentage of agreement between self-testing and testing by CHW in both countries. In Lesotho there was 97.6% (404/414, 95% CI: 95.6-99.8) agreement and in Zambia 99.8% (1116/1118, 95% CI: 99.4-100).

**Fig 1.**
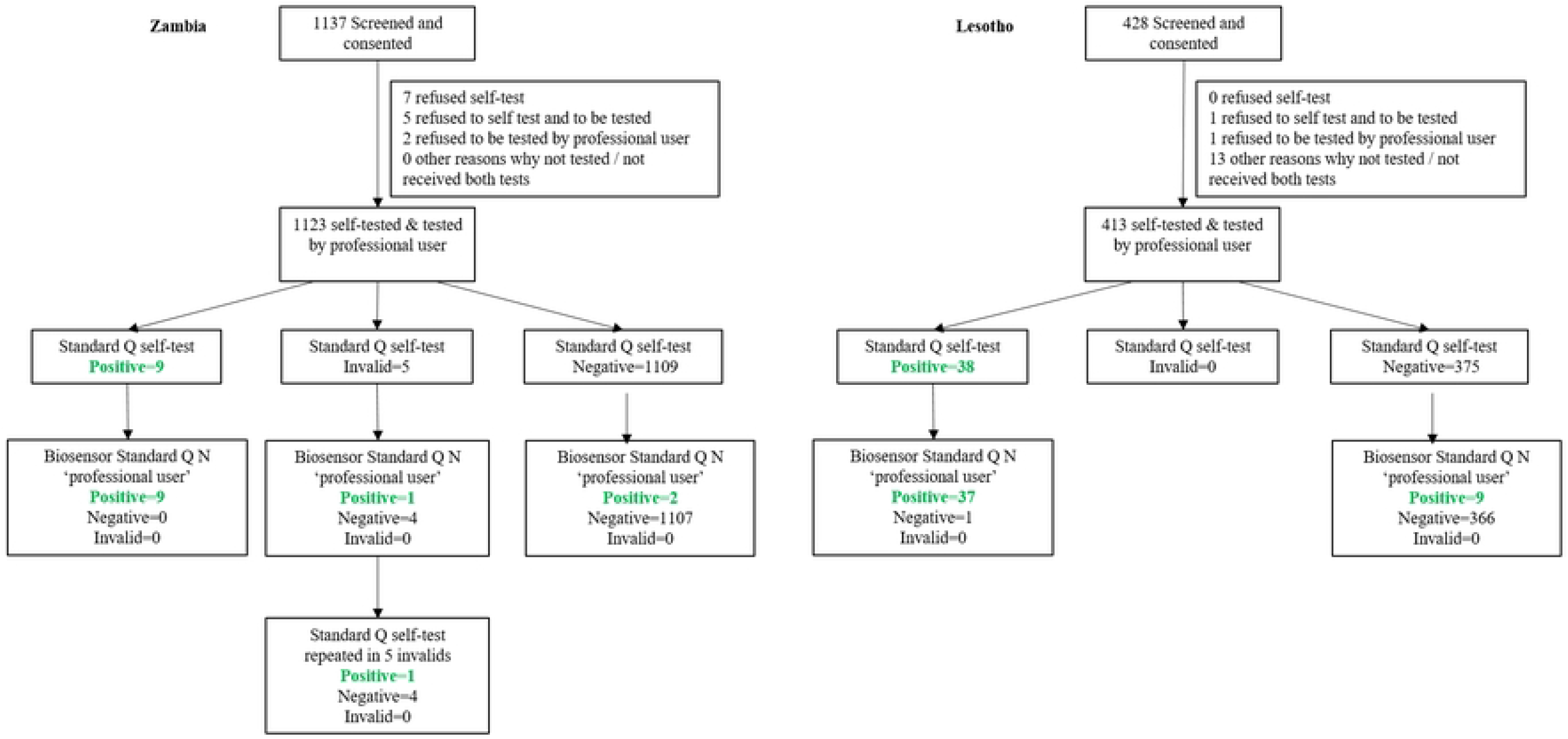
Study Flow chart in Zambia and Lesotho (STARD 2015)

**Table 6.** Percentage agreement between self-testing and testing by professional user using Ag-RDTs in Lesotho and Zambia.

| Country | Percent agreement between self-testing and testing by professional user using Ag-RDTs |  |  |
| --- | --- | --- | --- |
|  | n/N | % | 95% CI |
| **Lesotho** | 404/414 | 97.6 | 95.6-99.8 |
| **Zambia** | 1,116/1118 | 99.8 | 99.4-100 |

## Discussion

In this study, we found that despite differences in procedures and populations, the vast majority of participants in Zambia and Lesotho were able to understand and follow instructions for use and self-tested correctly. This resulted in a high level of agreement between Ag-RDT self-testing and testing by CHW in Lesotho (97.6%, 95% CI: 95.6-99.8%) and in Zambia (99.8%, 95% CI: 99.4-100%). Additionally, high acceptability of self-testing and a good usability was shown in Zambia. This is among the first studies to assess agreement between Ag-RDT self-testing and Ag-RDT testing by CHW, acceptability and usability of SARS-Cov-2 self-testing in the community in SSA.

Most of the research on self-testing actually assessed self-sampling and not the complete self-testing process. Such studies indicated that high quality samples can be taken by users, with self-taken samples tested against PCR yielding sensitivities and specificities close to, or above the WHO’s target product profile recommended sensitivity and specificity thresholds of 80% and 97% (19-24, 26-28). Regarding the complete self-testing process (i.e. self-collection and processing of sample and execution and interpretation of the self-test); some studies report good performance of self-testing against PCR(24, 29, 30), although others have found higher performance may be achieved when testing is conducted by trained personnel (24, 31).

Most of the previously mentioned studies come from high- and middle-income countries and/or from health care facility settings. There is only one other study to our knowledge that took place in the community in SSA and directly compared Ag-RDT self-testing against CHW Ag-RDT testing. This work from Sibanda et al., published in the WHO Interim Guidance on COVID-19 self-testing, presented findings from Malawi and Zimbabwe, although information on population, setting and procedures were not very detailed. This study reported high levels of agreement (97.9-100%) between self-testing and testing by health care workers in both rural and urban settings, using nasal Ag-RDTs, consistently with what was found in our study (3, 32). Both our study and the work of Sibanda et al., directly tested for operator effects while holding constant other factors that could influence test agreement or test accuracy (e.g. swab type, test type). It shows that self-testing in SSA can be performed accurately in the community and that with both supervised (Lesotho) and unsupervised self-testing (Zambia), high levels of agreement with CHW Ag-RDT testing can be achieved.

In Zambia, in terms of usability, most participants indicated that Ag-RDT self-testing using the manufacturer’s IFU was ‘very easy’ to perform. Pictures were the most useful piece of information in the IFU, according to almost all participants and most relied on the pictures rather than on the text for conducting the self-testing. This is also supported by findings from cognitive interviews in both countries, in which most participants said they used the pictures to follow the steps in the IFU’s. However, when participants were observed in Zambia, a considerable number of participants had difficulties following some of the steps correctly. Some of these difficulties such as removing the swab from the tube and extracting the liquid from the swab were also observed to be difficult for participants in the study in Malawi and Zimbabwe assessing usability of the same tests [23]. The study population in Malawi and Zimbabwe included health workers, who can be expected to be familiar with the testing process and able to understand instruction and to self-test easily compared to other populations. As evidenced with self-testing of HIV, difficulties following IFU for self-testing may vary depending on the population assessed. A study in Malawi and Zambia showed that performance of HIV self-testing can depend on literacy level and previous exposure to testing of the participant (33). Age has also been listed as a factor limiting the appropriate performance of self-testing (29).

According to cognitive interviews, participants of the study in Malawi and Zimbabwe thought that there were no major suggestions for change of IFU. In our study, cognitive interviews revealed that participants’ understanding and performance of some instructions was limited because of unclear images, use of difficult words and terms and the fact that some items were not included in the test kit package. A systematic review on the reliability of HIV rapid diagnostic tests for self-testing compared with testing by health-care workers concluded that most invalid results and errors in performance were related to user errors and manufacturing defects and could be mitigated with clarity on IFU (34). Complexity of the instructions can increase the possibility of failure in performance and incorrect interpretation of a result. Instructions could be adapted and validated for the cultural context and for less-skilled users, such as individuals with low literacy or visual impairments, including translation in local languages, clear and large print IFU, detailed images and descriptions, or electronic documents or audio instructions. Contrarily to what was done in Zambia in, Lesotho IFU were adapted and self-testing was guided. In Zambia, there were 5 participants that obtained an invalid result in the self-testing, no invalid results were found when tested by the professional user. Unfortunately, we do not have information on invalid results from Lesotho, limiting our ability to conclude whether guided self-testing with adaptation of the IFU’s result in better testing outcomes.

Regarding acceptability, in Zambia the refusal rate was very low (1%) and most participants indicated that they would be willing to conduct a self-test in the future and would recommend self-testing to a friend or family member. This observation is consistent with other studies showing high acceptability of self-tests for COVID-19 (8-14) and for other diseases (7). Studies on acceptability and willingness to self-test in LMIC showed that the general population, health workers, civil society and potential implementers, are supportive of the use of self-tests quite consistently across different countries (13).

The strengths of this study in Lesotho and Zambia include that the project was conducted in a community setting with real-world quantitative and qualitative data collection and analysis in two countries in SSA. In addition, the studies differed slightly in terms of study design and population, enabling us to demonstrate the utility of Ag-RDT self-testing in two settings in SSA. There were several limitations. First, the project was conducted during periods of low COVID-19 positivity rate in both countries. Further research is needed to assess if similar levels of agreement can be achieved during times of high transmission. Another weakness is that in Lesotho, details on invalid results were not recorded; tests were redone in case of invalid results and only the final result was documented. This could overestimate the level of agreement in the Lesotho data. A third limitation is that refusal in Lesotho was not documented consistently and may have led to an overestimation of acceptability in Lesotho.

## Conclusion

Our work indicates that Ag-RDT self-testing can be accurately performed, is acceptable and easy-to-use in SSA. These findings are relevant for pandemic preparedness and support the current recommendations from WHO and Africa CDC to include self-testing into COVID-19 testing strategies. We recommend further research on self-testing across different settings and populations and to assess cost-effectiveness to guide policy-making. Despite high agreement between self-testing and testing by community health workers, some participants experienced difficulties with certain steps in the IFU. We recommend manufacturers and/or health authorities to tailor IFU to local context and target population to improve user’s experience while self-testing and optimize the process.

## Data Availability

Data will be shared publicly but we still have not agreed where and, thus, we still do not have details of access

## Acknowledgements

The authors would like to express their appreciation and gratitude to all members of the study team in Zambia and Lesotho, and would like to acknowledge, in particular, all community and village health workers. We would also like to thank the Ministry of Health in both countries, and we appreciate the participation, time and information sharing of the study participants and their communities

## Supporting information

**S1 Appendix. Instructions for Use (IFU) for SARS-CoV-2 self-testing in Zambia**.

**S2 Appendix. Form used to fill in test results and ease-of-use questions in Zambia**.

**S3 Appendix. Instructions for Use (IFU) for SARS-CoV-2 self-testing in Lesotho**

**S4 Appendix. Definitions**

## Notes

### Competing Interest Statement

The authors have declared no competing interest.

### Funding Statement

This project is supported by a joint initiative between the Botnar Research Centre for Child Health and the European & Developing Countries Clinical Trials Partnership The funders had no role in study design, data collection and analysis, decision to publish, or preparation of the manuscript

